# Childhood trauma moderates schizotypy-related brain morphology: Analyses of 1,182 healthy individuals from the ENIGMA Schizotypy working group

**DOI:** 10.1101/2022.11.22.22282598

**Authors:** Yann Quidé, Oliver J. Watkeys, Emiliana Tonini, Dominik Grotegerd, Udo Dannlowski, Igor Nenadić, Tilo Kircher, Axel Krug, Tim Hahn, Susanne Meinert, Janik Goltermann, Marius Gruber, Frederike Stein, Katharina Brosch, Adrian Wroblewski, Florian Thomas-Odenthal, Paula Usemann, Benjamin Straube, Nina Alexander, Elisabeth J. Leehr, Jochen Bauer, Nils R. Winter, Lukas Fisch, Katharina Dohm, Wulf Rössler, Lukasz Smigielski, Pamela DeRosse, Ashley Moyett, Josselin Houenou, Marion Leboyer, James Gilleen, Sophia I. Thomopoulos, Paul M. Thompson, André Aleman, Gemma Modinos, Melissa J. Green

## Abstract

Schizotypy represents an index of psychosis-proneness in the general population often associated with childhood trauma exposure. Both schizotypy and childhood trauma are linked to structural brain alterations, and it is possible that trauma exposure moderates the extent of brain morphological differences associated with schizotypy. We addressed this question using data from a total of 1,182 healthy adults (age range: 18-65 years old, 647 females/535 males), pooled from nine sites worldwide, contributing to the Enhancing NeuroImaging Genetics through Meta-Analysis (ENIGMA) Schizotypy working group. All participants completed both the Schizotypal Personality Questionnaire Brief version (SPQ-B), and the Childhood Trauma Questionnaire (CTQ), and underwent a 3D T1-weighted brain MRI scan from which regional indices of subcortical grey matter volume and cortical thickness were determined. A series of multiple linear regressions revealed that differences in cortical thickness in four regions-of-interest were significantly associated with interactions between schizotypy and trauma; subsequent moderation analyses indicated that increasing levels of schizotypy were associated with thicker left caudal anterior cingulate gyrus, right middle temporal gyrus and insula, and thinner left caudal middle frontal gyrus, in people exposed to higher (but not low or average) levels of childhood trauma. This was found in the context of thicker bilateral medial orbitofrontal gyri, right rostral anterior cingulate gyrus, left temporal pole, left insula, and thinner left paracentral lobule directly associated with increasing levels of schizotypy. In addition, thinner left postcentral, superior parietal and lingual gyri, as well as thicker left caudal middle frontal gyrus and smaller left thalamus and right caudate were associated with increasing levels of childhood trauma exposure. These results suggest that alterations in brain regions critical for higher cognitive and integrative processes that are associated with schizotypy may be enhanced in individuals exposed to high levels of trauma.

## INTRODUCTION

Schizotypy refers to a set of behaviours, experiences, personality traits and beliefs, each normally distributed in the general population, that together reflect a continuum of psychosis-proneness.^1, 2^ Psychosis risk is influenced by environmental factors such as childhood trauma,^3, 4^ with exposure to abuse and/or neglect in childhood recognised as a major risk factor for psychotic disorders.^5, 6^ The concept of schizotypy is a useful construct for studying risk for schizophrenia in the general population ^7-9 10^ that avoids the potential confounding effects of clinical factors such as illness duration or medication dosage.^11^ Both schizotypy and childhood trauma exposure have been separately associated with overlapping changes in brain morphology,^12, 13^ and childhood trauma is associated with higher levels of schizotypy in people with a psychosis spectrum disorder (e.g., bipolar I disorder, schizophrenia) and in psychiatrically healthy individuals.^3, 4^ It is therefore possible that the differences in brain morphology associated with increasing schizotypy might be modified by exposure to childhood trauma, such that trauma exposure could moderate the relationship between schizotypy and variation in brain morphology. In fact, exposure to childhood trauma may influence the neurodevelopmental trajectories of brain regions critical for adequate affective and cognitive processes of vulnerable persons.

While structural brain abnormalities are ubiquitous among schizophrenia patients, the patterns of morphological differences associated with this illness are heterogeneous,^14^ just as schizotypy has been associated with inconsistent variation of subcortical volumes and/or cortical thickness.^12^ A recent meta-analysis from the Enhancing NeuroImaging Genetics through Meta-Analysis (ENIGMA) Schizotypy working group has identified that higher schizotypy scores, assessed using different questionnaires, were associated with thicker right medial orbitofrontal/ventromedial prefrontal cortex.^15^ In studies which used only the Schizotypal Personality Questionnaire (SPQ),^11, 16^ increasing schizotypy levels have been inconsistently associated with smaller thalamus^17^ and larger striatum (pallidum/putamen),^18^ as well as thinner *pars orbitalis*, rostral middle frontal and parahippocampal gyri^19^ and thicker right dorsolateral prefrontal cortex and dorsal premotor cortex/frontal eye fields.^17^

Exposure to childhood trauma is also associated with brain morphology variations, especially in stress-sensitive limbic regions, such as the hippocampus.^13, 20-23^ In a study with healthy individuals,^24^ childhood trauma exposure was found to moderate associations between schizotypy and grey matter covariation in a network including striatal and limbic regions. In that study, increased levels of schizotypy were associated with decreased grey matter volume in these regions, only in individuals who reported no childhood trauma exposure, but not for those who were exposed to childhood trauma.^24^ However, due to the large number of voxels included in whole-brain analyses, identified networks of brain regions largely overstep anatomical boundaries compared to clearly delineated regions-of-interest. It is thus possible that the use of whole-brain analyses may not strongly detect effects specific to individual brain regions.

Using harmonized data pooled from nine individual studies of healthy individuals expressing varying levels of schizotypy, contributing to the ENIGMA Schizotypy working group, we aimed to determine whether associations between schizotypy and indices of subcortical volume and cortical thickness are moderated by exposure to childhood trauma measured along a continuum of severity (ranging from absence to very severe). Analyses were conducted to examine interactions between childhood trauma severity and schizotypy in relation to variation in brain morphology (subcortical volumes, cortical thickness), especially in striatal, orbitofrontal/ventromedial and inferior prefrontal,^18, 25^ and stress-sensitive limbic (hippocampus, insula), frontal and striatal (caudate, putamen) regions^23^. We hypothesised that morphology of the subcortical (hippocampus, striatum) and cortical (insula, middle frontal gyrus) stress-sensitive regions would be differentially associated with schizotypy in individuals exposed to more severe levels of childhood trauma, compared to those who were not exposed. We specifically expected that more pronounced decreases in volume or thickness would reflect the additive effects of schizotypy and childhood trauma on these regions.

## METHODS

### Cohorts

Participants were 1,182 psychiatrically healthy individuals aged between 18 and 65 years [mean age=34.13 years old, standard deviation (SD)=12.35, N=647 (55%) females], pooled from nine worldwide cross-sectional cohorts contributing to the ENIGMA Schizotypy working group (see **Supplementary Table 1**). Data from each cohort was collected with participants’ written informed consent and ethical approval from local institutional review boards.

### Schizotypy

Schizotypy was measured using either the 74-items self-report SPQ^11^ or its brief 22-items version (SPQ-B).^16^ Only common items pertaining to the SPQ-B were used to calculate schizotypy indices to maintain consistency of measurement across sites. The total schizotypy score using the 22 items from the SPQ-B was calculated and used for focal analyses (score range: 0-22). Within the present dataset, the average SPQ total score was 3.27 (SD=3.35), ranging from 0 to 21. The distribution of the SPQ scores is presented in **Supplementary Figure 1**.

### Childhood trauma

The Childhood Trauma Questionnaire (CTQ)^26^ is a self-report measure of retrospective childhood trauma spanning domains of emotional, physical, and sexual abuse, as well as physical and emotional neglect. A score for each domain is calculated from five items on a 5-point Likert scale ranging from 1 (never true) to 5 (very often true). The total CTQ score (combining all types of trauma; score range: 25-125) was used for focal analyses. Within the present dataset, the average CTQ total score was 33.36 (SD=9.03), ranging from 25 to 84. The distribution of the CTQ scores is presented in **Supplementary Figure 2**.

### Brain imaging

All sites locally processed 3D T1-weighted structural brain MRI scans using FreeSurfer.^27, 28^ Following established ENIGMA protocols (http://enigma.usc.edu/protocols/imaging-protocols), grey matter volumes were extracted for 16 subcortical regions-of-interest (ROIs) from the automated segmentation (*aseg*) atlas (left and right lateral ventricle, thalamus, caudate, putamen, pallidum, accumbens, hippocampus and amygdala) and 68 cortical ROIs from the Desikan-Killiany atlas,^29^ as well as total intracranial volume (TIV) and mean cortical thickness (MThickness). Quality assessment was performed at each site prior to analyses, following the ENIGMA Quality assessment protocol <u>(ht</u>t<u>p://enigma.ini.usc.edu/protocols/imaging-protocols).</u> **Supplementary Table 2** summarises magnetic resonance imaging scanner manufacturers, models, magnet strengths, acquisition sequences, and the FreeSurfer version used to process the data at each site. Before entering statistical analyses, all neuroimaging variables (all subcortical and cortical ROIs, TIV, MThickness) were adjusted for scanning site using the modified empirical Bayes method ComBat for ENIGMA,^30^ as part of the R package ‘*sva’*.^31^

### Statistical analyses

Focal analyses were performed using R (v4.1.0)^32^ and RStudio (v1.3.1093). ^33^ Using a series of multiple linear regressions, the main effects of schizotypy (SPQ total score), childhood trauma severity (CTQ total score) and their interaction (the product of the mean-centred SPQ total score and the mean-centred CTQ total score) on indices of brain morphology in each ROI were separately determined. In case of significant interactions, moderation analyses were performed using the ‘*interactions’* R package.^34^ The moderation analyses tested the effects of schizotypy as the independent variable at three levels of the moderator: 1 SD below the mean CTQ score (low CTQ score), mean CTQ score, and 1 SD above the mean CTQ score (high CTQ score).^35^ In addition to the stress-sensitive regions hypothesized to be associated with the additive effects of schizotypy and trauma (i.e., hippocampus, striatum, insula, middle frontal gyrus), analyses were performed on all subcortical and cortical ROIs, for sake of completeness. For each set of analyses, only models surviving Bonferroni correction to account for the number of models tested (that is the number of ROIs) were considered (subcortical: *p*=0.05/16=3.13×10^−3^; cortical: *p*=0.05/68=7.35×10^−4^). The Davidson-McKinnon correction (HC3) was used to account for potential issues related to heteroskedasticity^36^ using the R package ‘*sandwich’*.^37, 38^ Within each significant model, statistical significance was set at a threshold of *p*<0.05. Age, quadratic age (age^2^, to model non-linear associations with age), sex and TIV (subcortical) or Mthickness (cortical) were entered as covariates in all analyses.

## RESULTS

Statistical details of all models are presented in **Table 1** for subcortical regions and **Table 2** for cortical regions.

**Table 1.** Results of the moderation analyses for all subcortical ROIs

| ROI | Model |  |  |  | Schizotypy (SPQ Total score) |  |  |  |  |  | Trauma (CTQ total score) |  |  |  |  |  | Schizotypy x Trauma |  |  |  |  |  |
| --- | --- | --- | --- | --- | --- | --- | --- | --- | --- | --- | --- | --- | --- | --- | --- | --- | --- | --- | --- | --- | --- | --- |
|  | *R*<sup>2</sup> | *F* | *df* | *p*-value | *b* | *se* | LLCI | ULCI | *t*-value | *p*-value | *b* | *se* | LLCI | ULCI | *t*-value | *p*-value | *b* | *se* | LLCI | ULCI | *t*-value | *p*-value |
| LLatVent | 0.236 | 53.001 | 8, 1174 | <3.13e<sup>-3</sup> | 25.8157 | 36.0018 | -44.8195 | 96.4508 | 0.7171 | 0.4735 | -4.2041 | 3.1185 | -10.3226 | 1.9143 | -1.3481 | 0.1779 | -4.2041 | 3.1185 | -10.3226 | 1.9143 | -1.3481 | 0.1779 |
| RLatVent | 0.223 | 49.454 | 8, 1174 | <3.13e<sup>-3</sup> | 16.5896 | 37.3549 | -56.7001 | 89.8793 | 0.4441 | 0.6570 | -10.4645 | 13.5707 | -37.0901 | 16.1611 | -0.7711 | 0.4408 | -3.5734 | 2.7701 | -9.0082 | 1.8615 | -1.2900 | 0.1973 |
| LThal | 0.468 | 148.078 | 8, 1165 | <3.13e<sup>-3</sup> | -2.1892 | 6.4770 | -14.8970 | 10.5187 | -0.3380 | 0.7354 | **-4.1407** | **2.1065** | **-8.2738** | **-0.0077** | **-1.9657** | **0.0496** | 0.0825 | 0.5279 | -0.9532 | 1.1182 | 0.1563 | 0.8758 |
| RThal | 0.555 | 210.669 | 8, 1170 | <3.13e<sup>-3</sup> | -1.3220 | 4.8072 | -10.7538 | 8.1098 | -0.2750 | 0.7834 | -2.7367 | 1.6126 | -5.9006 | 0.4272 | -1.6971 | 0.0899 | -0.0057 | 0.3837 | -0.7584 | 0.7471 | -0.0148 | 0.9882 |
| LCaud | 0.367 | 98.072 | 8, 1166 | <3.13e<sup>-3</sup> | 5.4770 | 3.7750 | -1.9296 | 12.8836 | 1.4508 | 0.1471 | -2.0742 | 1.3102 | -4.6448 | 0.4965 | -1.5831 | 0.1137 | -0.4659 | 0.3602 | -1.1726 | 0.2407 | -1.2937 | 0.1960 |
| RCaud | 0.409 | 117.071 | 8, 1169 | <3.13e<sup>-3</sup> | 3.4049 | 3.7816 | -4.0146 | 10.8243 | 0.9004 | 0.3681 | **-2.8040** | **1.2307** | **-5.2186** | **-0.3894** | **-2.2784** | **0.0229** | -0.4283 | 0.3304 | -1.0766 | 0.2200 | -1.2962 | 0.1952 |
| LPut | 0.410 | 114.581 | 8, 1136 | <3.13e<sup>-3</sup> | -0.7945 | 5.2990 | -11.1914 | 9.6025 | -0.1499 | 0.8808 | -0.3455 | 1.9399 | -4.1516 | 3.4607 | -0.1781 | 0.8587 | -0.2897 | 0.4126 | -1.0993 | 0.5198 | -0.7022 | 0.4827 |
| RPut | 0.490 | 161.354 | 8, 1159 | <3.13e<sup>-3</sup> | -2.7102 | 4.2757 | -11.0992 | 5.6789 | -0.6338 | 0.5263 | 0.8297 | 1.6271 | -2.3626 | 4.0220 | 0.5099 | 0.6102 | -0.6646 | 0.3550 | -1.3610 | 0.0319 | -1.8722 | 0.0614 |
| LPall | 0.189 | 36.409 | 8, 1059 | <3.13e<sup>-3</sup> | -3.5274 | 2.2886 | -8.0182 | 0.9633 | -1.5413 | 0.1235 | 0.2323 | 0.8798 | -1.4940 | 1.9585 | 0.2640 | 0.7918 | -0.3271 | 0.1897 | -0.6993 | 0.0451 | -1.7243 | 0.0849 |
| RPall | 0.204 | 43.551 | 8, 1154 | <3.13e<sup>-3</sup> | -0.5577 | 2.0439 | -4.5679 | 3.4525 | -0.2729 | 0.7850 | -0.3081 | 0.7648 | -1.8087 | 1.1924 | -0.4029 | 0.6871 | -0.2196 | 0.1657 | -0.5446 | 0.1054 | -1.3256 | 0.1853 |
| LHipp | 0.366 | 97.422 | 8, 1163 | <3.13e<sup>-3</sup> | 1.4600 | 3.0363 | -4.4973 | 7.4173 | 0.4808 | 0.6307 | -1.9439 | 1.0048 | -3.9153 | 0.0276 | -1.9345 | 0.0533 | 0.2464 | 0.2471 | -0.2384 | 0.7313 | 0.9973 | 0.3188 |
| RHipp | 0.355 | 93.082 | 8, 1166 | <3.13e<sup>-3</sup> | 3.6912 | 3.1805 | -2.5489 | 9.9313 | 1.1606 | 0.2461 | -2.0392 | 1.0987 | -4.1949 | 0.1166 | -1.8559 | 0.0637 | 0.1589 | 0.2500 | -0.3316 | 0.6495 | 0.6358 | 0.5250 |
| LAmyg | 0.323 | 80.772 | 8, 1161 | <3.13e<sup>-3</sup> | 1.6087 | 1.6142 | -1.5584 | 4.7758 | 0.9966 | 0.3192 | -0.8644 | 0.6379 | -2.1160 | 0.3872 | -1.3551 | 0.1757 | -0.0212 | 0.1413 | -0.2984 | 0.2559 | -0.1502 | 0.8806 |
| RAmyg | 0.324 | 80.752 | 8, 1156 | <3.13e<sup>-3</sup> | 1.6603 | 1.6700 | -1.6164 | 4.9369 | 0.9942 | 0.3204 | -0.5215 | 0.7413 | -1.9758 | 0.9329 | -0.7035 | 0.4819 | -0.0891 | 0.1678 | -0.4183 | 0.2401 | -0.5309 | 0.5956 |
| LAcc | 0.278 | 64.786 | 8, 1154 | <3.13e<sup>-3</sup> | -0.1146 | 0.9525 | -1.9834 | 1.7541 | -0.1204 | 0.9042 | -0.3938 | 0.3767 | -1.1328 | 0.3453 | -1.0454 | 0.2961 | 0.0179 | 0.0782 | -0.1355 | 0.1712 | 0.2288 | 0.8191 |
| RAcc | 0.362 | 94.826 | 8, 1152 | <3.13e<sup>-3</sup> | 0.1107 | 0.7542 | -1.3690 | 1.5904 | 0.1468 | 0.8833 | -0.0766 | 0.2957 | -0.6567 | 0.5035 | -0.2591 | 0.7956 | 0.0629 | 0.0674 | -0.0693 | 0.1952 | 0.9340 | 0.3505 |

ROI: region of interest; SPQ: schizotypal personality; CTQ: childhood trauma questionnaire; LLatVent: left lateral ventricle; RLatVent: right lateral ventricle; LThal: left thalamus; RThal: right thalamus; LCaud: left caudate; RCaud: right caudate; LPut: left putamen; RPut: right putamen; LPal: left pallidum; RPal: right pallidum; LHipp: left hippocampus; RHipp: right hippocampus; LAmyg: left amygdala; RAmyg: right amygdala; LAcc: left accumbens; RAcc: right accumbens; Adj *R*<sup>2</sup>: adjusted coefficient of determination; *se*: standard error; LLCI: bootstrapped 95% lower levels confidence interval; ULCI: bootstrapped 95% upper levels confidence interval
Statistically significant associations (*p*<0.05 within each model) are in bold and highlighted in grey

**Table 2.**
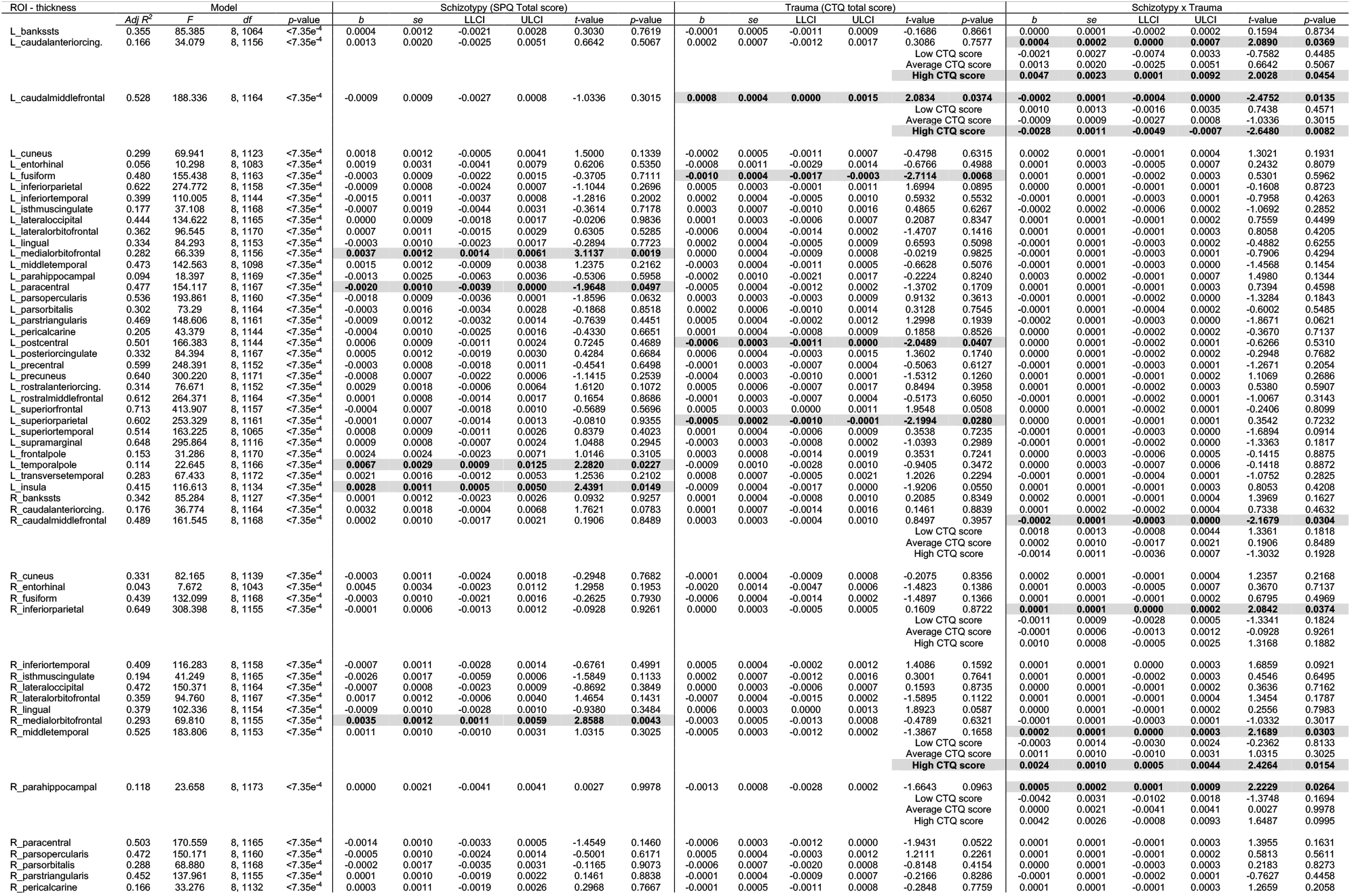
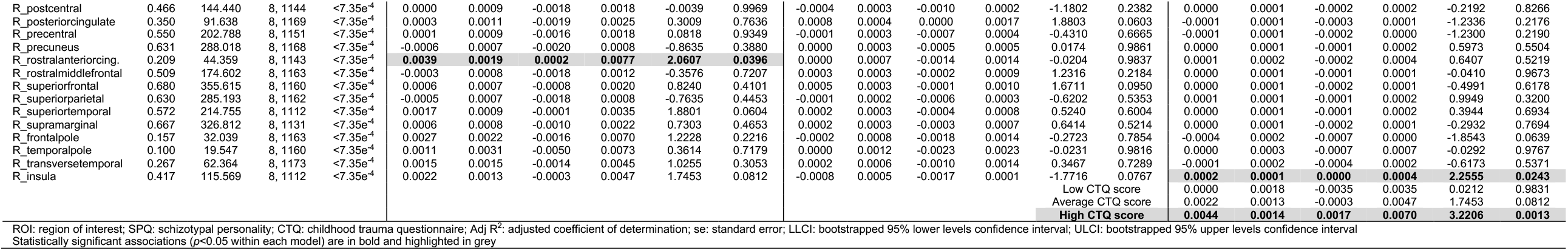
Results of the moderation analyses for all cortical ROIs

### Subcortical volume

All regression models were significant (all *p*<3.13×10^−3^; see **Table 1**). Main effects of trauma were evident in two ROIs, which showed a negative relationship with trauma severity: that is, greater severity of childhood trauma was significantly associated with lower volumes of the right caudate and left thalamus (see **Figure 1**). There were no other significant effects of trauma on other subcortical ROIs, and no significant main effects of schizotypy, or schizotypy-by-trauma interactions.

**Figure 1.**
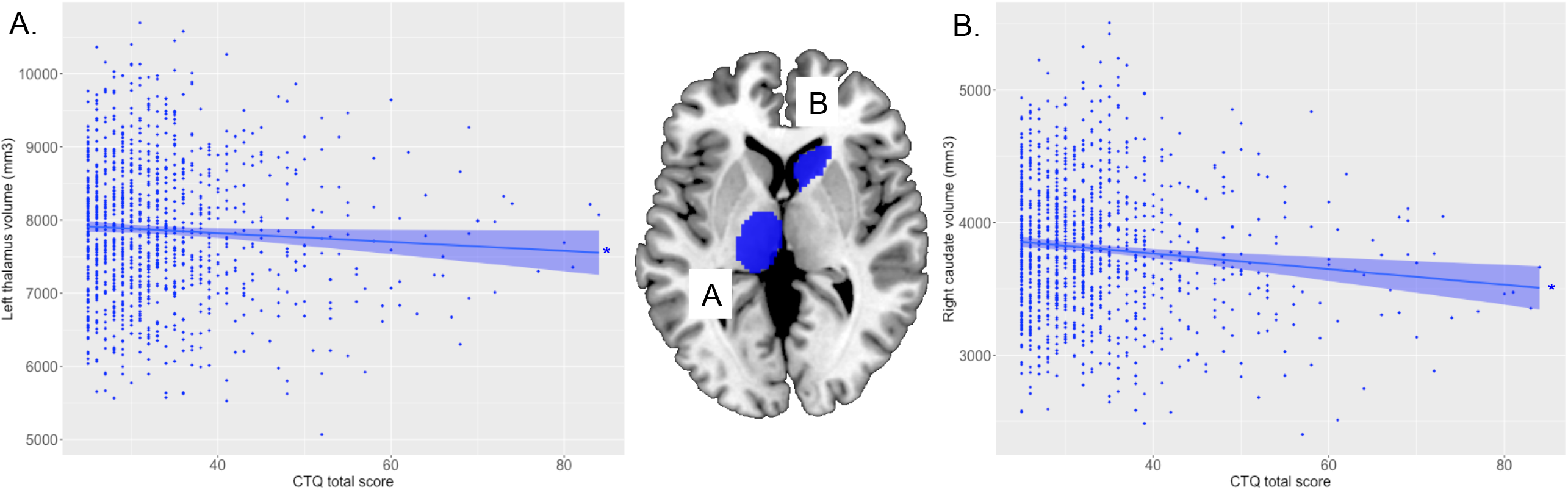
Relationship between severity of childhood trauma exposure and subcortical volumes. The severity of childhood trauma (CTQ total score) was associated with smaller volume of the (A) left thalamus and (B) right caudate. Coloured bands represent 95% confidence intervals, blue colour indicates negative associations. * *p*<0.05

### Cortical thickness

All regression models were significant (all *p*<7.35×10^−4^; see **Table 2**). The interaction between schizotypy and trauma severity was significantly associated with variation in the thickness of the left caudal anterior cingulate gyrus, the left and right caudal middle frontal gyri, the right inferior parietal gyrus, the right middle temporal gyrus, the right parahippocampal gyrus and the right insula. Moderation analyses indicated that higher levels of schizotypy were associated with thicker right middle temporal gyrus, left caudal anterior cingulate gyrus and right insula, as well as thinner left caudal middle frontal gyrus (see **Figure 2**), only among individuals exposed to high (but not low or average) trauma severity. Single slope analyses of the other models (right caudal middle frontal, inferior parietal and parahippocampal gyri) were not significant at any level of trauma severity.

**Figure 2.**
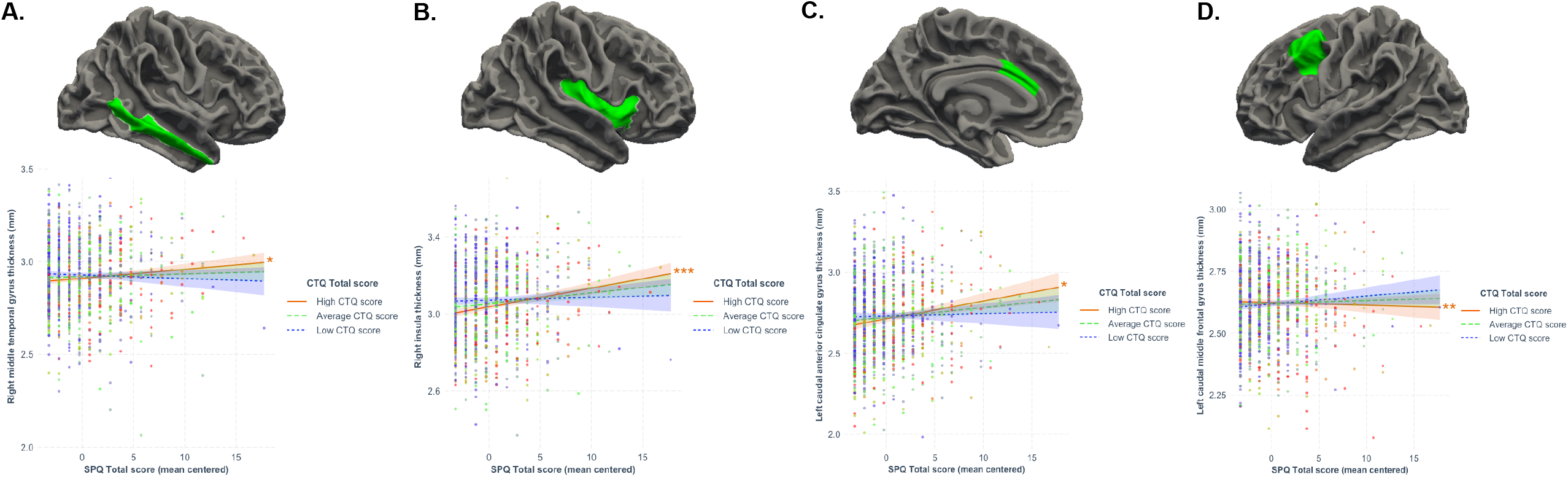
Results from the moderation analyses, following significant associations between the schizotypy-by-trauma interaction andcortical thickness. The severity of childhood trauma (CTQ Total score) moderated the relationship between schizotypy (SPQ total score) and thickness of the (A) right middle temporal gyrus, (B) right insula, (C) left caudal anterior cingulate gyrus, and (D) right caudal middle frontal gyrus. Moderation analyses indicated that increasing levels of schizotypy were significantly associated with thicker right middle temporal gyrus, insula and left caudal anterior cingulate gyrus, as well as with thinner left caudal middle frontal gyrus in individuals exposed to higher level (plain red lines), but not average (dashed green lines) or low (dotted blue lines) levels of trauma. Coloured bands represent 95% confidence intervals. * *p*<0.05; \*\**p*<0.01; \*\*\**p*<0.001

Main effects of schizotypy were also evident for six ROIs with higher levels of schizotypy significantly associated with thicker left and right medial orbitofrontal gyri, left temporal pole, left insula and right rostral anterior cingulate gyrus, as well as thinner left paracentral lobule (see **Figure 3**). In addition, main effects of trauma were evident for four cortical ROIs with greater severity of childhood trauma significantly associated with thicker left caudal middle frontal gyrus and thinner left fusiform, left posterior cingulate and left superior parietal gyri (see **Figure 4**).

**Figure 3.**
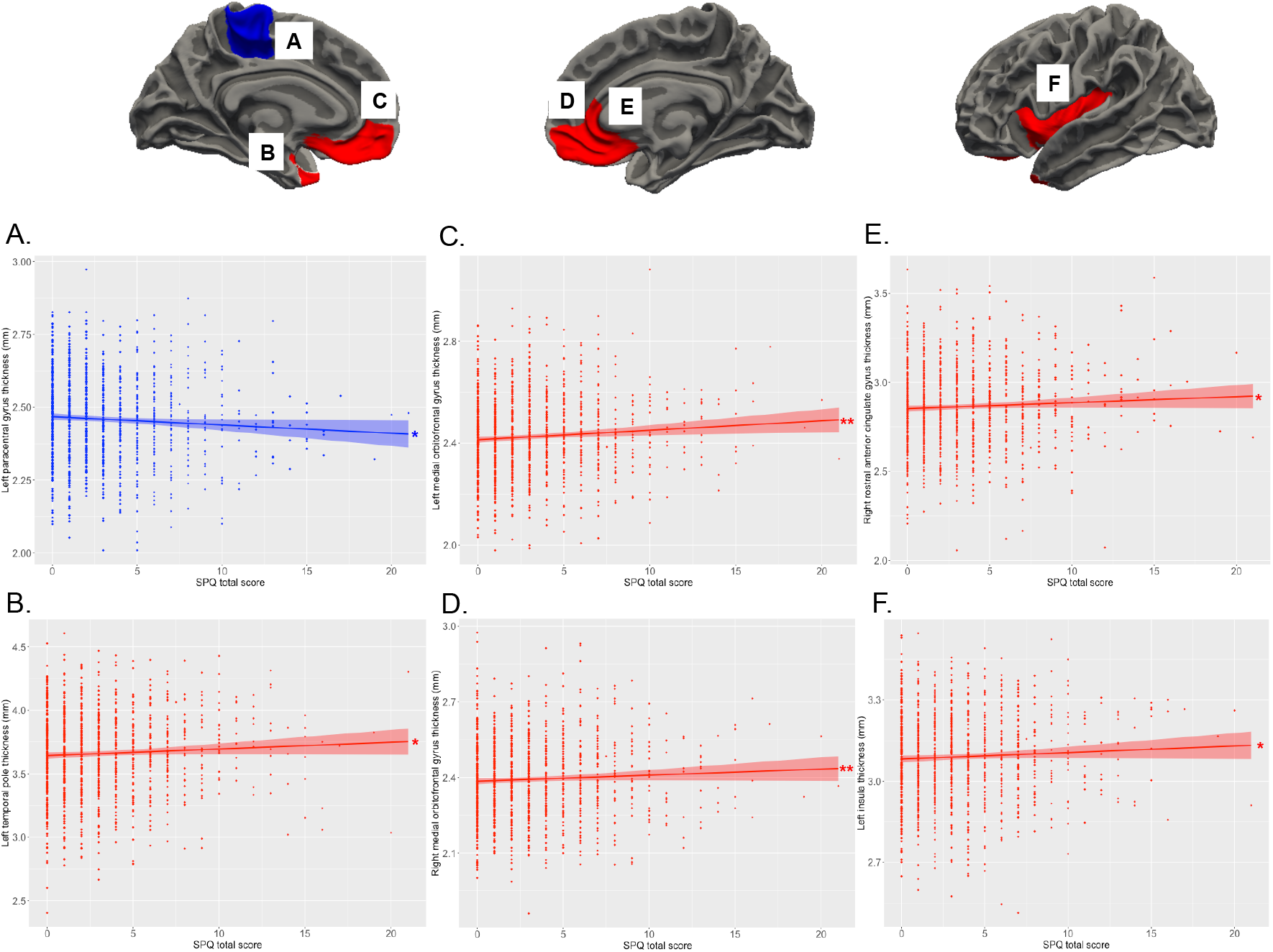
Relationship between the levels of schizotypy and cortical thickness. Increasing levels of schizotypy (SPQ total score) were associated with (A) thinner paracentral lobule, (B) thicker temporal pole, (C, D) thicker bilateral medial orbitofrontal gyri, (E) thicker right rostral anterior cingulate gyrus, and (F) thicker left insula. Coloured bands represent 95% confidence intervals, blue colour indicates negative associations, red colour indicates positive associations. * *p*<0.05; \*\**p*<0.01

**Figure 4.**
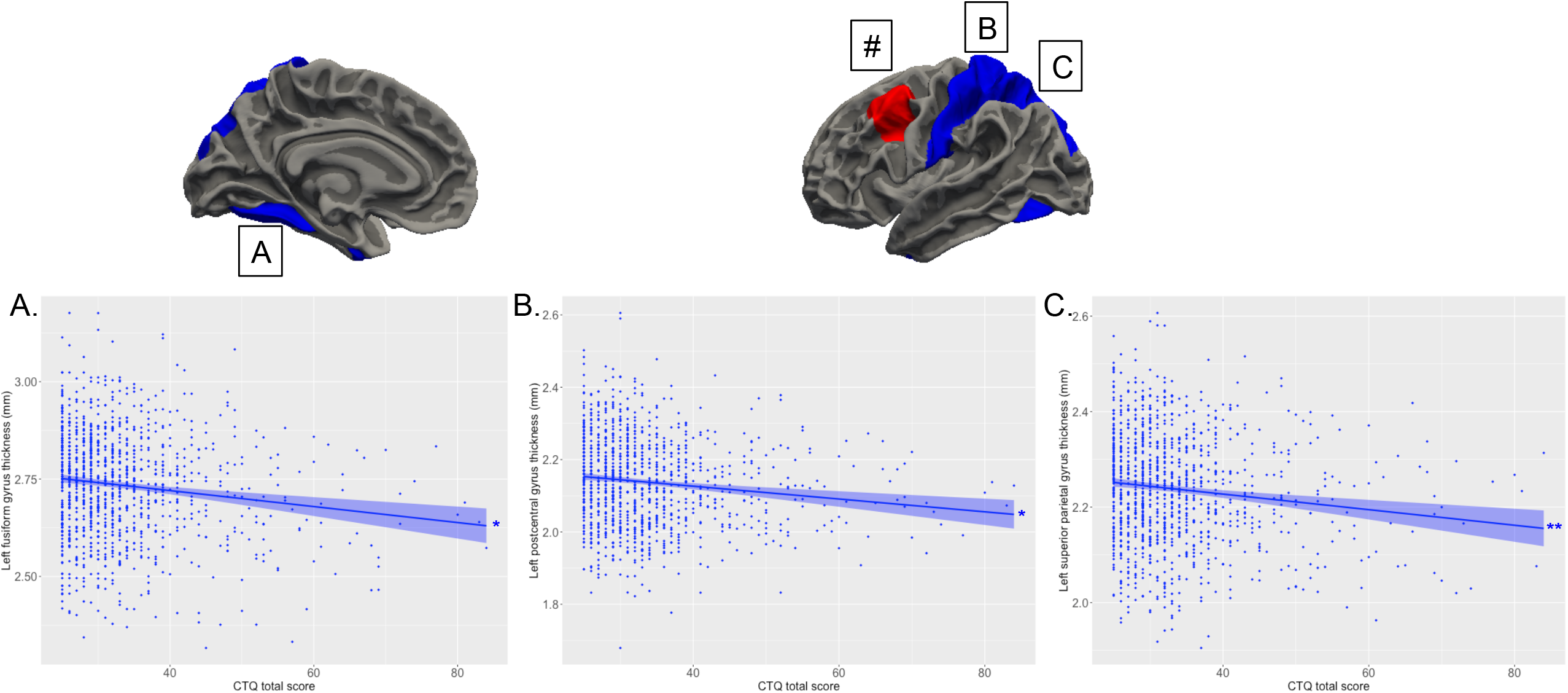
Relationship between the severity of childhood trauma exposure and cortical thickness. Increasing levels of childhood trauma (CTQ total score) were associated with thinner (A) fusiform gyrus, (B) left postcentral gyrus, (C) left superior parietal lobule. (#) The severity of childhood trauma was also associated with thicker left caudal middle frontal gyrus, but this association was confounded by a significant association between the schizotypy-by-trauma interaction indices of cortical thickness with this region (see Figure 2). Coloured bands represent 95% confidence intervals, blue colour indicates negative associations, red colour indicates positive associations. * *p*<0.05; \*\**p*<0.01

## DISCUSSION

These primary analyses of a large, pooled dataset assembled by the ENIGMA Schizotypy working group revealed that the severity of childhood trauma exposure moderated relationships between schizotypy and brain morphology in psychiatrically healthy adults. In particular, in individuals exposed to greater severity of childhood trauma, higher levels of schizotypy were associated with morphological differences in brain regions critical for higher cognitive processes such as executive functions (thinner caudal middle frontal, thicker caudal anterior cingulate gyri),^39^ exteroceptive, interoceptive (thicker insula)^40^ and multimodal integration (visual and auditory cortices, thicker middle temporal gyrus),^41^ as well as emotional processing (thicker caudal anterior cingulate gyrus).^42^ In addition, separate direct associations were evident between higher levels of schizotypy and brain morphology of affective (thicker medial orbitofrontal, rostral anterior cingulate gyri, temporal pole) and somatosensory regions (thinner left paracentral gyrus), as well as between the severity of childhood trauma and morphology of brain regions involved in somatosensory processing and integration (smaller thalamus, thinner postcentral gyrus), higher visual and cognitive functions, including reward and social processes (smaller caudate, thinner superior parietal, fusiform, and thicker rostral middle frontal gyri).

This large dataset revealed alterations in the thickness of cortical regions critical for cognitive and integrative processes in association with schizotypy, when more severe childhood trauma was experienced. These results indicate that the severity of childhood trauma experienced may impact the neurodevelopmental trajectories of these regions in relation to the expression of schizotypy. This is partially consistent with our prediction of additive effects of these two risk factors for psychosis on brain morphology: while decreases in volume or thickness were expected to reflect the deleterious additive effects of schizotypy and trauma on these stress-sensitive regions – which is the case for the left caudal middle frontal gyrus only – results show *increased* thickness associated with the additive effects of childhood trauma and schizotypy. Increased cortical thickness in multimodal integrative brain regions of people exposed to more severe childhood trauma may reflect a maladaptive integration of external and internal stimuli,^43^ evident in the expression of schizotypy. Indeed, alterations in brain morphology observed in individuals exposed to higher levels of trauma who did not report declared, full-blown psychiatric disorder at time of assessment, could either be associated with the presence of subclinical levels of psychiatric symptoms, or effective adaptation to trauma exposure.^13^ However, this interpretation is speculative and limited by the cross-sectional nature of the study, as it is possible that some individuals, in particular the younger participants included here, may have developed mental and/or physical health conditions associated with childhood trauma exposure at later stages of their life. In addition, aberrant development of these brain regions in association with schizotypy may reflect the influence of underlying genetic risk and other environmental factors not measured here,^44-46^ consistent with results found in clinical high-risk populations.^47, 48^ Future multimodal imaging-genetics studies, at different stages of neural development, are needed to confirm this interpretation.

The brain regions identified here as directly associated with schizotypy are largely consistent with our previous meta-analysis in 3,000 individuals,^15^ though not all of the associations reported here were significant in our previous meta-analysis (e.g., insula thickness). This is likely due to differences in the methods, as the former study was a meta-analysis of effects across a larger number of cohorts that used various scales to measure schizotypy (i.e., not limited to the SPQ-B, as in the present study). Importantly, there was no overlap between regions associated with schizotypy alone, and those associated with the schizotypy-by-trauma interaction. The regions associated solely with schizotypy are critical for affective integration,^49^ social cognition,^50^ and top-down emotional regulation,^51^ indicating that schizotypy may represent the long-term consequences of a maladaptive development of the affective brain. This is consistent with prior studies linking affective processes and schizotypy, even in the absence of diagnosed psychiatric conditions.^52^ Perhaps more surprisingly, schizotypy was associated with thinner paracentral lobule, a somatosensory region devoted to the representation of the lower limbs and genitals that could reflect movement and/or sexual dysfunctions for those reporting higher levels of schizotypy, as seen in schizophrenia.^53, 54^ This interpretation remains speculative, and in addition to including other imaging modalities, future studies using scales measuring indices of affective, mood or quality of life (e.g., anxiety, depression, motivation), are necessary to confirm this hypothesis.

Consistent with prior studies,^13, 23, 55, 56^ the present results also suggest that regions critical for somatosensory processes (smaller thalamus, thinner postcentral gyrus) and cognitive functions (smaller caudate, thinner superior parietal lobule, thicker caudal middle frontal gyrus), are sensitive to the severity of childhood trauma exposure. In particular, alterations of the somatosensory pathway may reflect long-term sensitization of this system following childhood trauma exposure, consistent with prior studies^57^. Smaller caudate may reflect aberrant reward sensitivity^58^ that may compromise striatal sensitivity to reward and other positive cues, previously found to be protective of the later development of trauma-related psychopathology.^59^ Similarly, and consistent with similar previous reports,^60^ childhood trauma may also have long-term consequences on the development of higher visual/attentional regions, as suggested in the present study by thinner fusiform gyrus and superior parietal lobule, respectively associated with the visual processing of “what” and “where”.^61^ Of note, thickness of the left caudal middle frontal gyrus was associated with both severity of trauma only (increased) and with schizotypy (decreased) when the severity of childhood trauma reported was higher, confirming the additive effects of these risk factors on the development of this critical brain region for higher cognitive functions. There was no other overlap between regions associated with the severity of trauma alone, and those associated with the schizotypy-by-trauma interaction. Unlike the results of a number of prior studies, smaller hippocampal volumes were not directly associated with the severity of trauma exposure.^13, 62^ Although perhaps surprising, this is consistent with recent meta-analytic results specifically accounting for sex in the context of reported psychopathology,^63^ and with a recent study in adult females.^64^ Long-term changes in hippocampal volume are therefore likely to be associated with other factors occurring (e.g., other stressful/traumatic life events) or developing (e.g., state/trait psychopathology) following childhood trauma exposure, rather than with trauma exposure itself. Future studies are warranted to better understand the short and long-term neuroanatomical consequences and dynamic trajectories of trauma exposure when not confounded by other factors (e.g., age, sex, psychopathology, etc.).

Notably, due to large methodological differences, the present results cannot be directly compared to a previous study showing decreased covariation in grey matter volume within a network of subcortical and cortical limbic regions (including the insula) in association with increasing levels of schizotypy, among individuals who reported no exposure to childhood trauma; however, these prior findings deserve mention in relation to the present results.^24^ While the present study used ROI-based subcortical volumes and cortical thickness derived from FreeSurfer segmentation, the previous study focused on *covariation* of brain volumetric indices in networks derived from independent component analysis (ICA) performed on whole-brain voxel-based morphometry (VBM) data. In addition, the use of a binary categorical index reflecting exposure/non-exposure to childhood trauma in the previous study meant that the severity of trauma exposure was not investigated, in contrast to the present study where the use of continuous variables is arguably more powerful for moderation analyses. Future studies are warranted that combine continuous independent variables with univariate (VBM) and multivariate (ICA) whole-brain imaging dependent variables, to better understand the role of childhood trauma on schizotypy-related grey matter changes, to be extended to indices of functional and structural connectivity.

This study has important strengths, including harmonised measures of schizotypy, childhood trauma and brain imaging in a large international cohort of medication-naïve healthy participants, but also has several limitations. First, this study used cross-sectional data, which prevented the investigation of developmental changes in brain morphology over time, in relation to schizotypy and exposure to childhood trauma. Age is a critical factor in the measurement of schizotypy: younger people usually present higher levels of schizotypy than older adults, and a peak in the expression of schizotypy is usually observed in adolescence.^65^ Different patterns of brain morphology associated with schizotypy may thus be evident at different stages of brain development, as suggested in cohorts of adolescents.^25, 66^ Future longitudinal studies are warranted, of adolescents and youths transitioning to adulthood, when levels of schizotypy and risks for first-episode psychosis are higher. Second, the population included in this study was largely comprised of middle-aged adults (average age was 34 years old, ranging from 18 to 65 years old). Although all statistical models included age as a covariate, more subtle effects of schizotypy may be evident at different developmental stages, and closer in time to trauma exposure. Third, only the overall severity of childhood trauma was examined in this study. It is possible that the severity of specific types and combinations of abuse or neglect, and when they occur during the brain’s development, may have different direct and interactive impacts on brain morphology. Though a recent study with 643 participants (including people with recent onset psychosis and people at clinical high risk of developing psychosis, amongst others) did not find different patterns of childhood trauma to be associated with differences in brain structure,^67^ this needs further scrutiny. Fourth, the retrospective self-report of childhood trauma exposure may have introduced recall biases toward the memory of traumatic events; however, the CTQ shows good reliability even in severely ill psychiatric populations,^68^ and good similarity with administered interviews.^69^ A recent meta-analysis showed some agreements between prospective and retrospective assessments of childhood trauma.^70^ Another limitation of this study is the relative low severity of childhood trauma exposure (mean CTQ total score = 33.36; range 25-84) and schizotypy (mean SPQ-B total score = 3.27; range 0-21) reported by the participants. This indicates that only few participants experienced more severe forms of childhood trauma and schizoptypy, potentially reducing the detection of more subtle effects associated with severe forms of both childhood trauma and schizotypy. Finally, the present study focused on the interactions between overall severity of reported childhood trauma and schizotypy and did not investigate the potential interactive effects of subtypes of childhood trauma (emotional, physical, sexual abuse, or emotional, physical neglect) and domains of schizotypy (cognitive-perceptual, interpersonal and disorganized). Because of the limited range of severity observed in reported childhood trauma and levels of schizotypy in psychiatrically healthy individuals, future studies will be necessary with sufficient statistical power to detect these subtle subtype effects, to identify relationships between subtypes of trauma, domains of schizotypy and brain development.

In conclusion, this study provides evidence for a moderating role of childhood trauma exposure in the relationship between schizotypy and brain morphology (thickness of middle temporal, insula, caudal anterior cingulate gyrus and caudal middle frontal gyrus) in a large cohort of healthy adults. Future large-scale multimodal imaging (morphological and functional) studies are needed to better understand the functional implications of alterations in brain morphology associated with risk for psychosis in the general population, their developmental timing, and their implications for risk of other mental disorders beyond the schizophrenia spectrum.

## Data Availability

All data produced in the present study are available upon reasonable request to the authors

## ACKNOWLEDGEMENTS

The ENIGMA-Schizotypy working group gratefully acknowledges support from the NIH Big Data to Knowledge (BD2K) award (U54 EB020403 to Paul M. Thompson).

**<u>FOR2107 – Marburg:</u>** This work was funded by the German Research Foundation (DFG grant FOR2107, KI588/14-1 and FOR2107, KI588/14-2 to Tilo Kircher). Igor Nenadić was supported by Deutsche Forschungsgemeinschaft (DFG), grants NE2254/2-1, NE2254/3-1, NDE2254/4-1.

**<u>FOR2107 – Muenster:</u>** This work was funded by the German Research Foundation (DFG, grant FOR2107 DA1151/5-1 and DA1151/5-2 to Udo Dannlowski; SFB-TRR58, Projects C09 and Z02 to Udo Dannlowski) and the Interdisciplinary Centre for Clinical Research (IZKF) of the medical faculty of Münster (grant Dan3/012/17 to Udo Dannlowski). Tim Hahn was funded by the German Research Foundation (DFG grants HA7070/2-2, HA7070/3, HA7070/4).

**<u>IGP:</u>** The Imaging Genetics in Psychosis (IGP) study was funded by Project Grants from the Australian National Health and Medical Research council (NHMRC; #630471 and #1081603), and the Macquarie University’s Australian Research Council Centre of Excellence in Cognition and its Disorders (CE110001021) awarded to Melissa J. Green. Emiliana Tonini was supported by the Australian Government Research Training Program (RTP) Scholarship (administered by the University of New South Wales), and a supplementary scholarship administered by Neuroscience Research Australia (NeuRA).

**<u>Zurich:</u>** This study was financially supported by the Donald C. Cooper-Fonds.

**<u>Paris</u>**: This study was supported by the French ANR “Agence Nationale pour la Recherche” (ANR MNP VIP and Labex BioPsy).

**<u>London:</u>** This work was supported by a Brain and Behavior Research Foundation NARSAD Young Investigator Grant (#21200, Lieber Investigator) and a Sir Henry Dale Fellowship jointly funded by the Wellcome Trust and the Royal Society (#202397/Z/16/Z), both awarded to Gemma Modinos.

**<u>Roehampton:</u>** This work was supported by a British Academy grant awarded to Dr James Gilleen (#SG150457).

## REFERENCES

1. Claridge GS. Schizotypy: Implications for illness and health. Oxford University Press: Oxford, England UK, 1997, xii, 340pp.

2. Grant P, Green MJ, Mason OJ. Models of Schizotypy: The Importance of Conceptual Clarity. Schizophr Bull 2018; 44(Suppl 2): S556–S563.

3. Velikonja T, Fisher HL, Mason O, Johnson S. Childhood trauma and schizotypy: a systematic literature review. Psychol Med 2015; 45(5): 947–963.

4. Quidé Y, Cohen-Woods S, O’Reilly N, Carr VJ, Elzinga BM, Green MJ. Schizotypal personality traits and social cognition are associated with childhood trauma exposure. Br J Clin Psychol 2018; 57(4): 397–419.

5. McGrath JJ, Saha S, Lim CCW, Aguilar-Gaxiola S, Alonso J, Andrade LH et al. Trauma and psychotic experiences: transnational data from the World Mental Health Survey. Br J Psychiatry 2017; 211(6): 373–380.

6. Varese F, Smeets F, Drukker M, Lieverse R, Lataster T, Viechtbauer W et al. Childhood adversities increase the risk of psychosis: a meta-analysis of patient-control, prospective and cross-sectional cohort studies. Schizophr Bull 2012; 38(4): 661–671.

7. Meehl PE. Toward an integrated theory of schizotaxia, schizotypy, and schizophrenia. Journal of Personality Disorders 1990; 4(1): 1–99.

8. Claridge GS. Origins of Mental Illness. Blackwell: Oxford, 1985.

9. Eysenck HJ. Dimensions of Personality. Routledge & K. Paul, 1950.

10. Kwapil TR, Barrantes-Vidal N. Schizotypy: looking back and moving forward. Schizophr Bull 2015; 41 Suppl 2: S366–373.

11. Raine A. The SPQ: a scale for the assessment of schizotypal personality based on DSM-III-R criteria. Schizophr Bull 1991; 17(4): 555–564.

12. Tonini E, Quidé Y, Kaur M, Whitford TJ, Green MJ. Structural and functional neural correlates of schizotypy: A systematic review. Psychol Bull 2021; 147(8): 828–866.

13. Teicher MH, Samson JA, Anderson CM, Ohashi K. The effects of childhood maltreatment on brain structure, function and connectivity. Nat Rev Neurosci 2016; 17(10): 652–666.

14. Shepherd AM, Laurens KR, Matheson SL, Carr VJ, Green MJ. Systematic meta-review and quality assessment of the structural brain alterations in schizophrenia. Neuroscience & Biobehavioral Reviews 2012; 36(4): 1342–1356.

15. Kirschner M, Hodzic-Santor B, Antoniades M, Nenadic I, Kircher T, Krug A et al. Cortical and subcortical neuroanatomical signatures of schizotypy in 3004 individuals assessed in a worldwide ENIGMA study. Mol Psychiatry 2021.

16. Raine A, Benishay D. The SPQ-B: A brief screening instrument for schizotypal personality disorder. Journal of Personality Disorders 1995; 9(4): 346–355.

17. Kuhn S, Schubert F, Gallinat J. Higher prefrontal cortical thickness in high schizotypal personality trait. J Psychiatr Res 2012; 46(7): 960–965.

18. Meller T, Ettinger U, Grant P, Nenadic I. The association of striatal volume and positive schizotypy in healthy subjects: intelligence as a moderating factor. Psychol Med 2020; 50(14): 2355–2363.

19. DeRosse P, Nitzburg GC, Ikuta T, Peters BD, Malhotra AK, Szeszko PR. Evidence from structural and diffusion tensor imaging for frontotemporal deficits in psychometric schizotypy. Schizophr Bull 2015; 41(1): 104–114.

20. Hart H, Rubia K. Neuroimaging of child abuse: a critical review. Front Hum Neurosci 2012; 6: 52.

21. Teicher MH, Samson JA. Childhood maltreatment and psychopathology: A case for ecophenotypic variants as clinically and neurobiologically distinct subtypes. Am J Psychiatry 2013; 170(10): 1114–1133.

22. Lim L, Radua J, Rubia K. Gray matter abnormalities in childhood maltreatment: a voxel-wise meta-analysis. Am J Psychiatry 2014; 171(8): 854–863.

23. Popovic D, Ruef A, Dwyer DB, Antonucci LA, Eder J, Sanfelici R et al. Traces of Trauma: A Multivariate Pattern Analysis of Childhood Trauma, Brain Structure, and Clinical Phenotypes. Biol Psychiatry 2020; 88(11): 829–842.

24. Quidé Y, Tonini E, Watkeys OJ, Carr VJ, Green MJ. Schizotypy, childhood trauma and brain morphometry. Schizophr Res 2021; 238: 73–81.

25. Derome M, Zoller D, Modinos G, Schaer M, Eliez S, Debbane M. Developmental trajectories of subcortical structures in relation to dimensional schizotypy expression along adolescence. Schizophr Res 2020; 218: 76–84.

26. Bernstein DP, Stein JA, Newcomb MD, Walker E, Pogge D, Ahluvalia T et al. Development and validation of a brief screening version of the Childhood Trauma Questionnaire. Child Abuse and Neglect 2003; 27(2): 169–190.

27. Fischl B, Salat DH, Busa E, Albert M, Dieterich M, Haselgrove C et al. Whole brain segmentation: automated labeling of neuroanatomical structures in the human brain. Neuron 2002; 33(3): 341–355.

28. Fischl B, Salat DH, van der Kouwe AJW, Makris N, Segonne F, Quinn BT et al. Sequence-independent segmentation of magnetic resonance images. NeuroImage 2004; 23 Suppl 1: S69–84.

29. Desikan RS, Segonne F, Fischl B, Quinn BT, Dickerson BC, Blacker D et al. An automated labeling system for subdividing the human cerebral cortex on MRI scans into gyral based regions of interest. NeuroImage 2006; 31(3): 968–980.

30. Radua J, Vieta E, Shinohara R, Kochunov P, Quidé Y, Green MJ et al. Increased power by harmonizing structural MRI site differences with the ComBat batch adjustment method in ENIGMA. NeuroImage 2020; 218: 116956.

31. Leek JT, Johnson WE, Parker HS, Fertig EJ, Jaffe AE, Zhang Y et al. sva: Surrogate Variable Analysis. R package version 3.34.0. 2019.

32. R Core Team. R: A language and environment for statistical computing. R Foundation for Statistical Computing: Vienna, Austria, 2021.

33. RStudio Team. RStudio: Integrated Development Environment for R. In: RStudio P (ed). Boston, MA, USA, 2021.

34. Long JA. interactions: Comprehensive, User-Friendly Toolkit for Probing Interactions. R package version 1.1.5. 2021.

35. Cohen J, Cohen P, West SG, Aiken LS. Applied Multiple Regression/Correlation Analysis for the Behavioral Sciences. 3rd edn, 2003, online resource (735 pages).

36. Hayes AF, Cai L. Using heteroskedasticity-consistent standard error estimators in OLS regression: An introduction and software implementation. Behavior Research Methods 2007; 39(4): 709–722.

37. Zeileis A. Econometric Computing with HC and HAC Covariance Matrix Estimators. Journal of Statistical Software 2004; 11(10): 1–17.

38. Zeileis A, Köll S, Graham N. Various Versatile Variances: An Object-Oriented Implementation of Clustered Covariances in R. Journal of Statistical Software 2020; 95(1): 1–36.

39. Alvarez JA, Emory E. Executive function and the frontal lobes: a meta-analytic review. Neuropsychol Rev 2006; 16(1): 17–42.

40. Craig AD. How do you feel--now? The anterior insula and human awareness. Nature Reviews Neuroscience 2009; 10(1): 59–70.

41. Mesulam MM. From sensation to cognition. Brain 1998; 121 (Pt 6): 1013–1052.

42. Stevens FL, Hurley RA, Taber KH. Anterior cingulate cortex: unique role in cognition and emotion. J Neuropsychiatry Clin Neurosci 2011; 23(2): 121–125.

43. Harricharan S, McKinnon MC, Lanius RA. How Processing of Sensory Information From the Internal and External Worlds Shape the Perception and Engagement With the World in the Aftermath of Trauma: Implications for PTSD. Front Neurosci 2021; 15: 625490.

44. Newbury JB, Arseneault L, Caspi A, Moffitt TE, Odgers CL, Belsky DW et al. Association between genetic and socioenvironmental risk for schizophrenia during upbringing in a UK longitudinal cohort. Psychol Med 2020: 1–11.

45. Walter EE, Fernandez F, Snelling M, Barkus E. Genetic Consideration of Schizotypal Traits: A Review. Front Psychol 2016; 7: 1769.

46. Tonini E, Quidé Y, Whitford TJ, Green MJ. Cumulative sociodemographic disadvantage partially mediates associations between childhood trauma and schizotypy. Br J Clin Psychol 2021.

47. Andreou C, Borgwardt S. Structural and functional imaging markers for susceptibility to psychosis. Mol Psychiatry 2020; 25(11): 2773–2785.

48. de Wit S, Wierenga LM, Oranje B, Ziermans TB, Schothorst PF, van Engeland H et al. Brain development in adolescents at ultra-high risk for psychosis: Longitudinal changes related to resilience. Neuroimage Clin 2016; 12: 542–549.

49. Lieberman MD, Straccia MA, Meyer ML, D. M, Tan KM. Social, self, (situational), and affective processes in medial prefrontal cortex (MPFC): Causal, multivariate, and reverse inference evidence. Neurosci Biobehav Rev 2019; 99: 311–328.

50. Abu-Akel A, Shamay-Tsoory S. Neuroanatomical and neurochemical bases of theory of mind. Neuropsychologia 2011; 49(11): 2971–2984.

51. Motzkin JC, Philippi CL, Wolf RC, Baskaya MK, Koenigs M. Ventromedial prefrontal cortex is critical for the regulation of amygdala activity in humans. Biol Psychiatry 2015; 77(3): 276–284.

52. Lewandowski KE, Barrantes-Vidal N, Nelson-Gray RO, Clancy C, Kepley HO, Kwapil TR. Anxiety and depression symptoms in psychometrically identified schizotypy. Schizophr Res 2006; 83(2-3): 225–235.

53. Walther S, Strik W. Motor symptoms and schizophrenia. Neuropsychobiology 2012; 66(2): 77–92.

54. de Boer MK, Castelein S, Wiersma D, Schoevers RA, Knegtering H. The facts about sexual (Dys)function in schizophrenia: an overview of clinically relevant findings. Schizophr Bull 2015; 41(3): 674–686.

55. Edmiston EE, Wang F, Mazure CM, Guiney J, Sinha R, Mayes LC et al. Corticostriatal-limbic gray matter morphology in adolescents with self-reported exposure to childhood maltreatment. Arch Pediatr Adolesc Med 2011; 165(12): 1069–1077.

56. Baker LM, Williams LM, Korgaonkar MS, Cohen RA, Heaps JM, Paul RH. Impact of early vs. late childhood early life stress on brain morphometrics. Brain Imaging Behav 2013; 7(2): 196–203.

57. Cassiers LLM, Sabbe BGC, Schmaal L, Veltman DJ, Penninx B, Van Den Eede F. Structural and Functional Brain Abnormalities Associated With Exposure to Different Childhood Trauma Subtypes: A Systematic Review of Neuroimaging Findings. Front Psychiatry 2018; 9: 329.

58. Haber SN, Knutson B. The reward circuit: linking primate anatomy and human imaging. Neuropsychopharmacology 2010; 35(1): 4–26.

59. McLaughlin KA, Lambert HK. Child Trauma Exposure and Psychopathology: Mechanisms of Risk and Resilience. Curr Opin Psychol 2017; 14: 29–34.

60. Everaerd D, Klumpers F, Zwiers M, Guadalupe T, Franke B, van Oostrom I et al. Childhood abuse and deprivation are associated with distinct sex-dependent differences in brain morphology. Neuropsychopharmacology 2016; 41(7): 1716–1723.

61. Goodale MA, Westwood DA. An evolving view of duplex vision: separate but interacting cortical pathways for perception and action. Curr Opin Neurobiol 2004; 14(2): 203–211.

62. Dannlowski U, Stuhrmann A, Beutelmann V, Zwanzger P, Lenzen T, Grotegerd D et al. Limbic scars: long-term consequences of childhood maltreatment revealed by functional and structural magnetic resonance imaging. Biological Psychiatry 2012; 71(4): 286–293.

63. Calem M, Bromis K, McGuire P, Morgan C, Kempton MJ. Meta-analysis of associations between childhood adversity and hippocampus and amygdala volume in non-clinical and general population samples. Neuroimage Clin 2017; 14: 471–479.

64. Herzog JI, Thome J, Demirakca T, Koppe G, Ende G, Lis S et al. Influence of Severity of Type and Timing of Retrospectively Reported Childhood Maltreatment on Female Amygdala and Hippocampal Volume. Sci Rep 2020; 10(1): 1903.

65. Debbané M, Badoud D, Balanzin D, Eliez S. Broadly defined risk mental states during adolescence: disorganization mediates positive schizotypal expression. Schizophr Res 2013; 147(1): 153–156.

66. Derome M, Tonini E, Zoller D, Schaer M, Eliez S, Debbane M. Developmental Trajectories of Cortical Thickness in Relation to Schizotypy During Adolescence. Schizophr Bull 2020.

67. Haidl TK, Hedderich DM, Rosen M, Kaiser N, Seves M, Lichtenstein T et al. The non-specific nature of mental health and structural brain outcomes following childhood trauma. Psychol Med 2021: 1–10.

68. Fisher HL, Craig TK, Fearon P, Morgan K, Dazzan P, Lappin J et al. Reliability and comparability of psychosis patients’ retrospective reports of childhood abuse. Schizophr Bull 2011; 37(3): 546–553.

69. Gayer-Anderson C, Reininghaus U, Paetzold I, Hubbard K, Beards S, Mondelli V et al. A comparison between self-report and interviewer-rated retrospective reports of childhood abuse among individuals with first-episode psychosis and population-based controls. J Psychiatr Res 2020; 123: 145–150.

70. Baldwin JR, Reuben A, Newbury JB, Danese A. Agreement Between Prospective and Retrospective Measures of Childhood Maltreatment: A Systematic Review and Meta-analysis. JAMA Psychiatry 2019; 76(6): 584–593.

