## Supplementary for "Childhood trauma moderates schizotypy-related brain morphology: Analyses of 1,182 healthy individuals from the ENIGMA Schizotypy working group"

**– Supplementary Material –**

***Corresponding author**
Dr Yann Quidé, NeuroRecovery Research Hub, School of Psychology, Biological Sciences (Biolink) Building, UNSW Sydney, NSW, 2052, Australia.


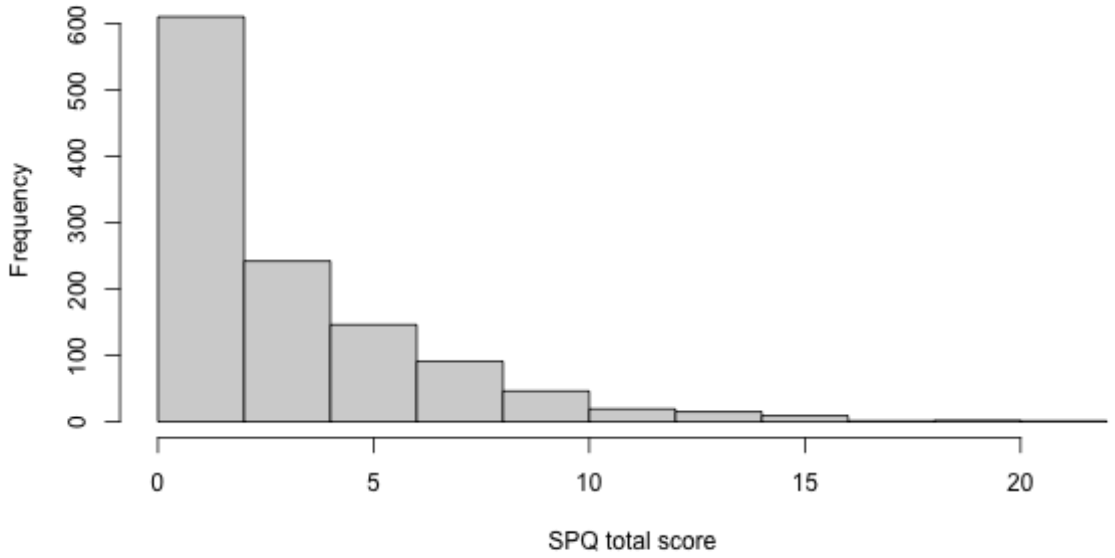


**Supplementary Figure 1.** Distribution of the SPQ total score (maximum possible range 0-22)

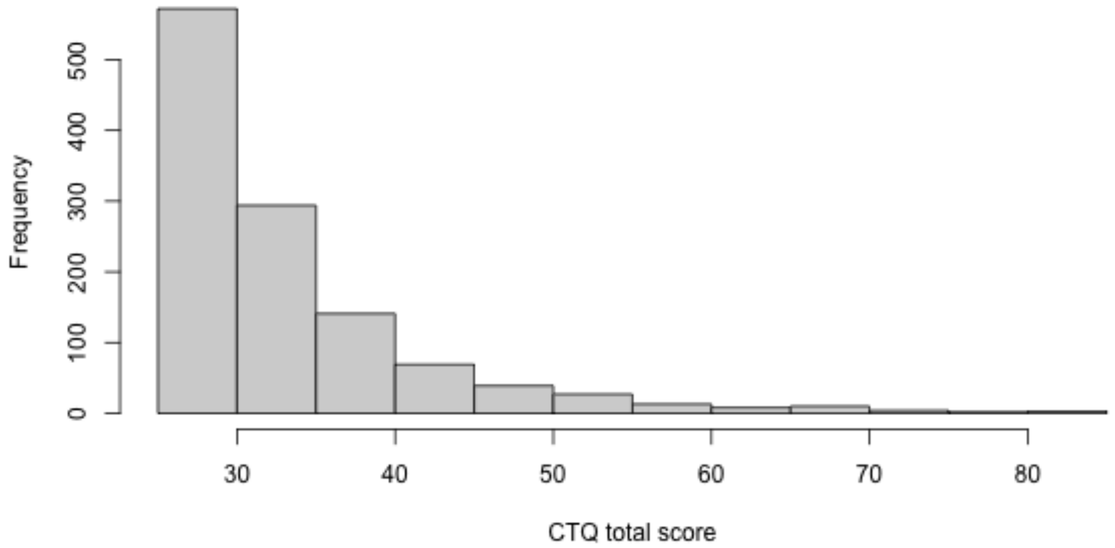


**Supplementary Figure 2.** Distribution of the CTQ total score (maximum possible range 25-125)

**Supplementary Table 1.** Sample sizes of the contributing sites

| Sites | Sample size (N) |
|---|---|
| FOR2107 – Marburg | 408 |
| FOR2107 – Muenster | 224 |
| Muenster Neuroimaging Cohort (MNC) | 169 |
| New York – Hillside | 165 |
| Zurich | 59 |
| IGP | 52 |
| London | 45 |
| Paris | 40 |
| Roehampton | 20 |

| **Supplementary Table 2.** Scanner details for each site | | | | | |
|---|---|---|---|---|---|
| Cohorts | Scanner type | Magnet strength | Acquisition sequence | Sequence parameters | FreeSurfer version |
| FOR2107 – Marburg | Siemens Magnetom TiroTim syngo | 3T | 3D-MPRAGE | TR 1900ms, TE 2.26ms, TI 900ms, FA 9°, voxel size 1.0x1.0x1.0mm³, Acquisition Direction Sagittal, 176 slices, slice gap 0.5mm. | 5.3 |
| FOR2107 – Muenster | Siemens Prisma | 3T | 3D-MPRAGE | TR 2130ms, TE 2.28ms, TI 900ms, FA 8°, voxel size 1.0x1.0x1.0mm³, Acquisition Direction Sagittal, 192 slices, no slice gap. | 5.3 |
| Muenster Neuroimaging Cohort (MNC) | Philips Gyroscan Intera | 3T | 3D-fast gradient echo sequence (turbo field echo) | TR 7.4ms, TE 3.4ms, FA 9°, two signal averages, inversion prepulse every 814.5 ms, acquired over a FOV of 256 (feet-head [FH]) × 204 (anterior-posterior [AP]) × 160 (right-left [RL]) mm, phase encoding in AP and RL direction, reconstructed to cubic voxels of .5 × .5 × .5 $mm^3$ | 5.3 |
| New York – Hillside | GE | 3T | 3D-SPGR | TR 7.5 ms, TE 3 ms, matrix 256x256, FOV 240 mm, 216 contiguous images, thickness 1mm, interleaved | 6.0 |
| IGP | Philips Achieva TX | 3T | 3D-MPRAGE | TR 8.9ms, TE 4.1ms, FOV 240mm, matrix 268 x 268, 200 sagittal slices, slice thickness 0.9mm (no gap) | 5.3 |
| Zurich | Philips Achieva | 3T | 3D-MPRAGE | TR 8.2ms; TE 3.8ms; FA 8°; voxel size, 1×1×1 $mm^3$; FOV 160 × 240 $mm^2$, 160 slices | 6.0 |
| London1b | Philips Intera | 3T | 3D fast-field echo (FFE) sequence | TR 25 ms, TE 4.6 ms, FOV 260 mm, matrix 256x256, 160 contiguous axial slices of 1-mm thickness, voxel size 1x1x1 $mm^3$ | 6.0 |
| Paris | Siemens Tim Trio | 3T | 3D-MPRAGE | TR 2300 ms, TE 2.98 ms, 160 slices ; voxel size, 1.0 × 1.0 × 1.1 | 5.3 |
| Roehampton | Siemens | 3T | 3D-MPRAGE | TR 2000ms, TE 2.07ms, FA 11°; voxel size, 1×1×1 $mm^3$; slice thickness 1mm; matrix 256x256, 176 slices256 | 6.0 |

TR: repetition time, TE= echo time, FA: flip angle; FOV: field of view
